# Transmission of B.1.617.2 Delta Variant between vaccinated healthcare workers

**DOI:** 10.1101/2021.11.19.21266406

**Authors:** Steven A. Kemp, Mark TK Cheng, Will Hamilton, Kimia Kamelian, INSACOG Consortium, Sujit Singh, Partha Rakshit, Anurag Aggrawal, Chris Illingworth, Ravindra K. Gupta

## Abstract

Breakthrough infections with SARS-CoV-2 Delta variant have been reported in vaccine recipients and in individuals infected with previous variants. However the potential for fully vaccinated individuals (two doses) to transmit SARS-CoV-2 is unclear. We here analyse data from health care workers in two hospitals in India, constructing probable transmission networks from epidemiological and virus genome sequence data using a suite of computational approaches. Among known cases we identify a high probability that doubly vaccinated individuals transmitted SARS-CoV-2, and potential cases of virus transmission between individuals who had received two doses of vaccine. Our findings highlight the need for ongoing infection control measures even in highly vaccinated populations.

## Introduction

The B.1.617.2 (Delta) variant is now the most common lineage circulating in India and worldwide. Vaccine efficacy (VE) for the Delta variant is estimated at around 50% for mild symptomatic infection after two doses of either mRNA or adenovirus-based vaccine platforms^1^. However, VE for moderately-severe and severe disease is estimated as 95%^1^. Breakthrough infections of the Delta variant have been reported in vaccine recipients, as well as re-infection of individuals infected with previously circulating variants^2-4^. We previously reported 155 occurrences of vaccine breakthrough infections^5^ amongst frontline Delhi healthcare workers (HCW), all of whom had one or two doses of the Covishield (ChAdOx1) vaccine, the majority of whom were vaccinated >21 days prior to symptomatic presentation.

Existing literature suggests that there are similar viral loads between vaccinated and unvaccinated^6,7^, but with steeper decay^8^ over time in vaccinated persons. Furthermore the probability of culturing virus appears lower in the vaccinated. It is unclear whether individuals with vaccine breakthrough are able to transmit to other vaccinated individuals, or whether transmission is driven solely by the unvaccinated. Here we utilise *in silico* approaches combining genomic and epidemiological investigation data to identify probable Delta variant transmission events between vaccinated HCW in Indian hsopitals.

## Results

### ChAdOx1 nCoV-19 Breakthrough Infection

All symptomatic HCWs in two hospitals who had received one or two doses of the ChAdOx1 nCoV-19 mRNA vaccine (AZD1222), in addition to a set of unvaccinated HCWs, underwent RT-PCR testing within one day of symptom onset as part of a hospital staff symptomatic testing program. From these cases we identified cases of breakthrough infection in staff who had received two doses of the vaccine. In hospital A, there were 81 breakthrough infections amongst 1100 HCWs, and in hospital B, 32 infections amongst 4000 HCWs, as previously reported^5^.

Among the 113 cases from hospitals A and B, 12.4% were administrative staff, 31.9% were nurses, 40.7% were primary physicians, 7.1% were paramedics and 3.5% were pharmacists. The remaining 3.5% of breakthrough infections consisted of medical interns (1.8%), and laboratory workers (1.8%) (**Supplementary Table 1**). The median interval between receiving a second vaccination and date of positive RT-PCR test was 45 days (range 3-78 days). Amongst the breakthrough infections of doubly-vaccinated HCW, 90.7% were infected with B.1.617.2, 5.3% by B.1.1.7-like, 1.3% by B.1.538. (**Supplementary Table 2**). The most commonly reported symptoms were fever (82.3% of all cases), cough (43.4%), myalgia (20.4%), and loss of smell/taste (14.2%) (**Supplementary Table 3**).

Whole genome sequencing was used to characterise staff nose and throat swab samples. In hospital A, 66 sequences with high quality whole genome coverage of >90% were generated, including 43 cases of breakthrough infection. In hospital B, high quality genome sequences were generated from 52 samples, including all 32 symptomatic breakthrough infections.

### Community Sequencing

Between April – May 2021, >99% of SARS-CoV-2 sequences in India were assigned Delta (B.1.616.2) lineages. However, since August 2021, the Delta sub-lineages have proliferated - predominantly AY.12 – across India and elsewhere^9^. To determine the relationship between community and HCW sequences, we inferred a maximum-likelihood phylogeny to estimate dispersion (**Supplementary Figure 1**). The analysis suggested multiple introductions into hospitals A and B, with subsequent intra-hospital transmission. We found significant partitioning of the inferred phylogeny, separating the sequences into disintict clades. Mutations relative to the Delta consensus that were found in these cases were spread across the SARS-CoV-2 genome. None of the identified SNPs were in homoplasic regions of the genome. We note that the individuals 115, 127, and 305 in hospital A had identical virus genomes.

### Linkage between vaccinated HCW

Considering cases of infection with the B.1.617.2 Delta variant, we inferred putative transmission pairs using the A2B-COVID^10^ software package, which identifies pairs of individuals for whom symptom onset dates and viral genome sequences are consistent with direct transmission. Using parameters inferred for the Delta SARS-CoV-2 lineage (**Supplementary Text, Supplementary Figure 2**) we identified 35 putative transmission events involving 14 HCWs in hospital A, and 26 putative events involving 13 patients in hospital B (**Supplementary Figures 3, 4**).

Data from these cases were then analysed with the A2B-Network software package^11^ which calculates the probability of specific networks of transmission events between a set of individuals. To reduce excessive computational load, potential transmission events in hospital A were filtered to remove those involving the gain of more than one SNP. From the remaining cases we identified a network involving six HCWs, two of whom had received their second dose of vaccine at least 14 days prior to reporting symptoms (**Figure 1**). A total of 1381 possible transmission networks between these individuals were identified, allowing us to calculate the probabilities of different sets of transmission events. Under the conditions of our model there was a 92.1% chance that one of the two individuals receiving a second vaccine 14 days prior to infection was the source of transmission to another individual in the network. If this criterion is relaxed to 7 days between second vaccination and infection, we estimated a 99.9% chance that at least one of the three such individuals infected another individual in the network, with a 98.7% probability of direct virus transmission between these three individuals. The filtering of this subset individuals from the larger network could not have increased any of these statistics (**Supplementary Figure 5**). Other unrelated networks in hospital A did not involve transmission from vaccinated HCWs to unvaccinated HCWs (**Supplementary Figure 6**).

**Figure 1.**
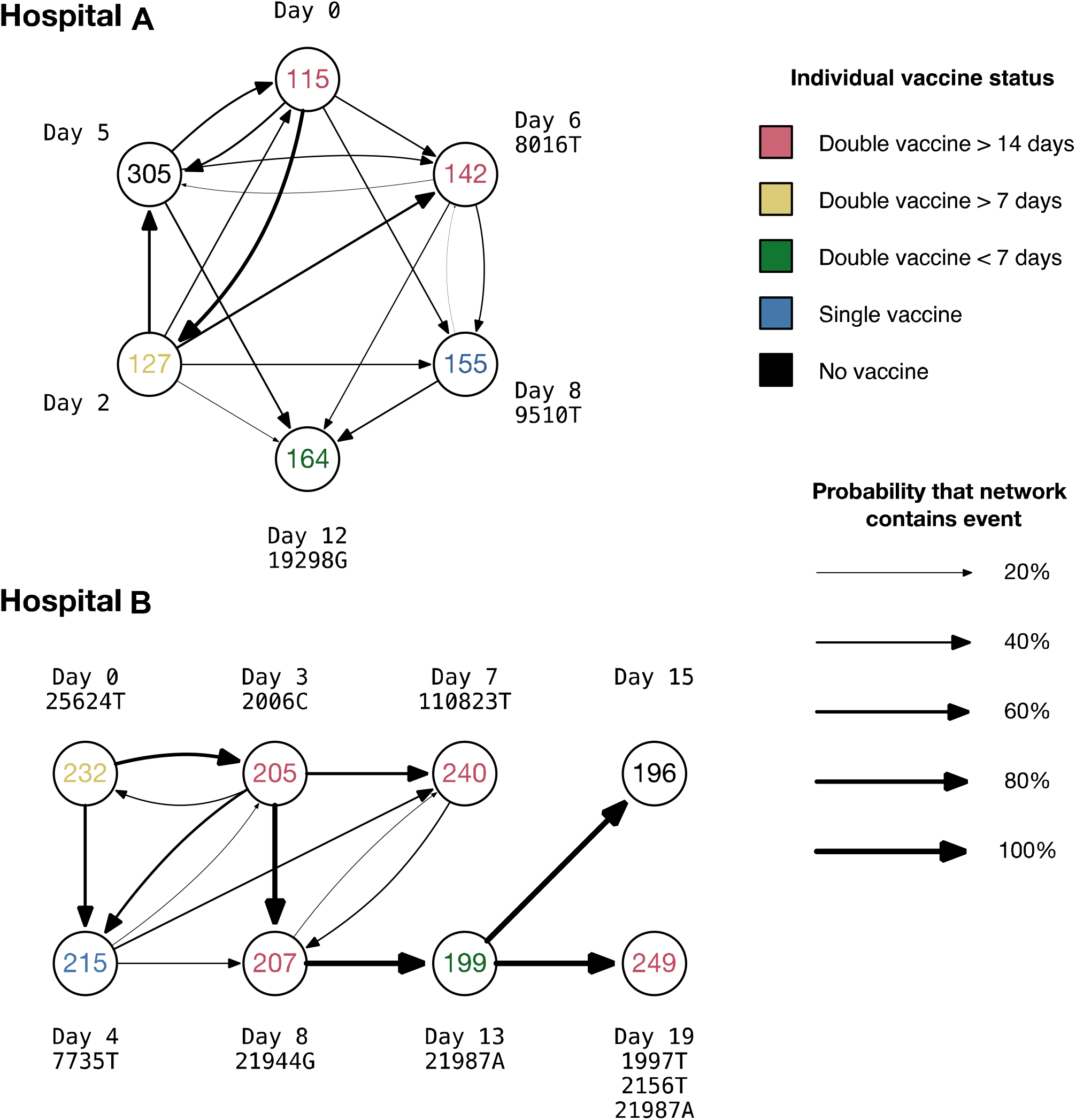
Potential transmission networks between HCWs. Individual labels are coloured according to vaccine status, including the timing prior to infection at which the second vaccine was given, where relevant. The thickness of lines between individuals show the probabilities of distinct pairwise transmission events between individuals; these probabilities are conditional on transmission having occurred between the individuals observed in each network. Labels show the relative dates on which individuals became symptomatic, and respective gains of nucleotides in sequences collected from each individual with respect to the mutual consensus.

Analysis of potential transmission events in hospital B identified a network involving eight HCWs, four of whom had received their second dose of vaccine at least 14 days prior to reporting symptoms (**Figure 1**). A total of 128 possible transmission networks between these individuals were identified, all of which implied that one of the four individuals who received a second vaccine at least 14 days prior to infection was the source of transmission to another individual in the network. Among the five individuals who were infected more than seven days post second vaccination, we found a 97.8% chance of a transmission event having occurred between individuals vaccinated with two doses.

While our analysis could not account for unobserved data, we performed validation calculations evaluating the self-consistency of the observed sequence data with the inferred patterns of virus transmission. Comparing our data against simulations suggested that the transmission network Inferred for hospital A is self-consistent with the observed sequence data, though the network inferred for hospital B is potentially distorted by missing data, with too high a number of individuals carrying unique mutations (**Supplementary Figure 7**). Details of simulations are provided in **Supplementary Text**.

### Transmission Networks

Inferred transmission networks involved a variety of medical staff. In hospital A, a common genome sequence was shared by the junior medical staff member P115, the nursing student P127, and by P305, a member of the nursing staff, who collectively became symptomatic over a period of six days. Three other staff had virus genomes which differed from this by a single SNP each. The ophthalmologic junior medical staff P142 became symptomatic a day after P305; the SNP 8016T causes a nonsynonymous change in ORF1ab, A2584V. P155 was also a member of the nursing staff; the SNP C9509T leads to the nonsynonymous substitution T3082I in ORF1ab. Finally, P164 was a paramedic; the SNP A19298G represents a nonsynonymous change in the ORF1ab (Y6345C).

In hospital B, the individuals P205, P207, and P240 all received their second dose of vaccine at least 14 days before testing positive. P205 was a paediatrician, P207 and P240 both adult physicians. Further details of symptoms, job titles, symptoms, and positive test dates, where known, are shown in **Supplementary Table 4**. Full details of SNPs are given in **Supplementary Figure 8 and 9**.

## Discussion

As the Delta variant dominates globally, information on the transmission amongst fully vaccinated individuals is needed to dictate the need for future infection control and vaccination boost strategies. Our study highlights the plausibility of Delta variant transmission in a healthcare setting from and between fully vaccinated individuals. Our study has caveats; firstly by only considering symptomatic cases, we exclude HCWs who are asymptomatic or fail to report their infection status. Secondly, the number of valid samples is further reduced by incomplete sampling and subsequent sequence filtering, likely resulting in incomplete transmission chains. Calculations suggested that the networks inferred for hospital B are likely to be affected by missing data, though the data from hospital A was not clearly affected.

Although vaccination is still highly effective for Delta in protecting against severe disease^12^, we have demonstrated that breakthrough infections still occur in healthcare settings in individuals within 60 days of the second dose when circulating neutralising antibody levels are at their highest^13^. Given the risk of onward transmission to potentially very vulnerable patients (including people in whom vaccination is less effective, such as those with compromised immune systems), our findings highlight the need for ongoing infection control measures even in highly vaccinated populations, and mask wearing in individuals who have received two doses of vaccine, in order to limit onward transmission.

## Data Availability

All data produced are available online at https://github.com/TKMarkCheng/Indian_HCW

## Methods

### Study Design

From 25th March – 10th May 2021 all frontline healthcare workers who presented with symptoms consistent with SARS-CoV-2 were diagnostically tested for the presence of SARS-CoV-2 by means of reverse transcriptase polymerase chain reaction (RT-PCR) using TRUENAT or CBNAAT (GENEXPERT). Findings were expressed as the cycle threshold (Ct) for the gene encoding the nucleocapsid protein (N gene) for hospital A, and the gene encoding for the envelope protein (E gene) for hospital V. A Ct value of less than 30 was considered to be infective. A vaccine breakthrough infection was defined as detection of SARS-CoV-2 in a sample collected from an individual 21 days after receipt of a second dose of ChAdOx1.

Ethical approval for the study of vaccine elicited antibodies in sera from vaccinees was obtained from the East of England Cambridge Central Research Ethics Committee Cambridge (REC ref. 17/EE/0025). Use of convalescent sera had ethical approval from the South Central Berkshire B Research Ethics Committee (REC ref. 20/SC/0206; IRAS 283805). Studies involving HCWs (including testing and sequencing of respiratory samples) were reviewed and approved by The Institutional Human Ethics Committees of the National Centre for Disease Control (NCDC) and Council Of Scientific And Industrial Research–Institute Of Genomics And Integrative Biology (CSIR IGIB) (NCDC/2020/NERC/14 and CSIRIGIB/IHEC/202021/01). Participants provided informed consent.

### Bioinformatics and Phylogenetic analysis

Fasta consensus sequences were obtained from two separate Hospitals in Delhi, India. All sequences were concatenated into a single fasta and aligned to reference strain MN908947.3 with mafft v7.847^14^ using the --keeplength and --addfragments options. Following this, all sequences were screened for number of gaps and N-regions using the Nextclade v1.5.4 (https://clades.nextstrain.org/) server. All sequences were assigned a lineage with Pangolin v3.1.11^15^, pangoLEARN (dated 9^th^ August 2021) and scorpio v0.0.14. Sequences that could not be assigned a lineage were discarded. After assigning lineages, all sequences with more than 5% N-regions were also discarded.

To contextualise outbreak sequences, all sequences from India with lineage defined as B.1.617.2 from the month of April 2021 were downloaded from the Global Initiative on Sharing Avian Influenza Data (GISAID) EpiCoV database. Incomplete (<29,000 base pair), duplicate, and low-quality sequences (defined as equal to or more than 5% Ns; less than 95% genome coverage) were excluded.

### SNP distance, variant calling and annotation

Single-nucleotide polymorphisms (SNPs), relative to the Delta variant strain, were identified by re-alignment of groupings of sequences based on each hospital to the Delta variant consensus reference (MZ359841.1) using mafft v7.8477 with the --keeplength and --addfragments parameters. Initial analysis of pairwise SNP distance between each patient was conducted using snp-dists v0.8.2 with default parameters.

Following this, a VCF of acquired mutations of each patient with respect to the reference strain MN908947.3 (Wuhan-Hu-1) is calculated by snp-sites v2.5.1^16^ using the -v and -c option. Multiallelic variants are broken down into biallelic variant representations and are subsequently annotated by snpEff v5.0e^17^ with reference to MN908947.3. The nucleotide and amino acid variants between each transmission pairs are extracted from the VCF, and SNP number verified using an in-house script (https://github.com/TKMarkCheng/Indian_HCW). The VCFs were manually scanned to identify homoplasic/problematic sites (https://github.com/W-L/ProblematicSites_SARS-CoV2).

### Estimating likelihood of person-to-person transmission

As an initial assessment of whether participants in the study had passed an infection to another, we utilised A2B-COVID^10^. This software considers data from individuals in a pairwise fashion, considering the timing of symptom onset and virus genome sequences in order to assess for each pair of individuals A and B whether the data are consistent with an underlying model of direct virus transmission from A to B. Data from each pair are described as ‘consistent’ with transmission, ‘borderline’, or ‘unlikely’ to have been produced given direct transmission.

Network reconstruction was performed using the A2B-Network software^11^ Again using dates of symptom onset and virus genome sequences, this identifies plausible networks via which all of the individuals in a set could have transmitted the virus between themselves, and assesses the probability of each such network having occurred, given our model assumptions. In this manner, the code produces ensembles of networks, describing the extent to which the data constrain the possible routes of transmission of the virus. The probabilities we report were calculated by summing network likelihoods. For example, the probability that a network contains a transmission event between doubly vaccinated individuals is the sum of the probabilities of the networks which contain at least one such transmission event. Our model assumes a set of underlying parameters describing the transmission dynamics and rate of evolution of the SARS-CoV-2 virus. The network probabilities we report are conditional on these parameters and upon the assumption that the individuals to whom we apply the model are connected by a transmission network.

### Network validation

Transmission networks were validated for self-consistency using a model of simulated SARS-CoV-2 outbreaks. Simulations used identical parameters to the network inference model to describe the transmission dynamics and rate of evolution of the SARS-CoV-2 virus. Via repeated simulation, we evaluated whether the genetic properties of the virus genome sequences in our datasets, described in terms of the number of unique mutations observed, and the number of mutations unique to each individual, were consistent with what we would expect to observe if the networks we had inferred were complete and correct. Further details are given in **Supplementary Text**.

## Notes

### Competing Interest Statement

The authors have declared no competing interest.

### Funding Statement

R.K.G. is supported by a Wellcome Trust Senior Fellowship in Clinical Science (WT108082AIA). This study was supported by the Cambridge NIHRB Biomedical Research Centre. S.A.K. is supported by the Bill and Melinda Gates Foundation via PANGEA grant OPP1175094.

### Author Declarations

Ethical approval for the study of vaccine elicited antibodies in sera from vaccinees was obtained from the East of England Cambridge Central Research Ethics Committee Cambridge (REC ref. 17/EE/0025). Use of convalescent sera had ethical approval from the South Central Berkshire B Research Ethics Committee (REC ref. 20/SC/0206; IRAS 283805). Studies involving HCWs (including testing and sequencing of respiratory samples) were reviewed and approved by The Institutional Human Ethics Committees of the National Centre for Disease Control (NCDC) and CSIR IGIB(NCDC/2020/NERC/14 and CSIRIGIB/IHEC/202021/01). Participants provided informed consent.

