## Supplementary material for "Transmission of B.1.617.2 Delta Variant between vaccinated healthcare workers": supp figures

### Slide 1
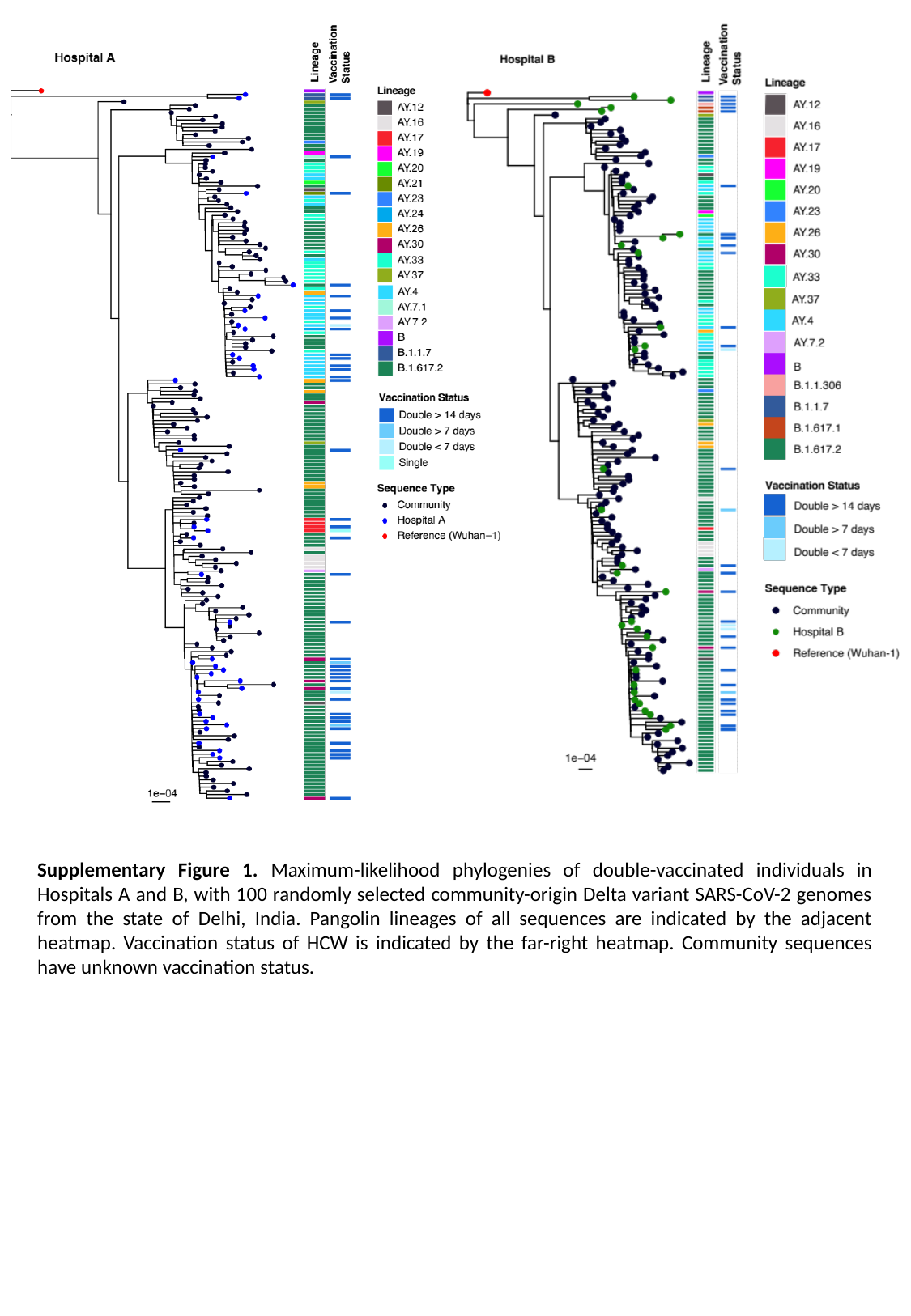

Supplementary Figure 1. Maximum-likelihood phylogenies of double-vaccinated individuals in Hospitals A and B, with 100 randomly selected community-origin Delta variant SARS-CoV-2 genomes from the state of Delhi, India. Pangolin lineages of all sequences are indicated by the adjacent heatmap. Vaccination status of HCW is indicated by the far-right heatmap. Community sequences have unknown vaccination status.

### Slide 2
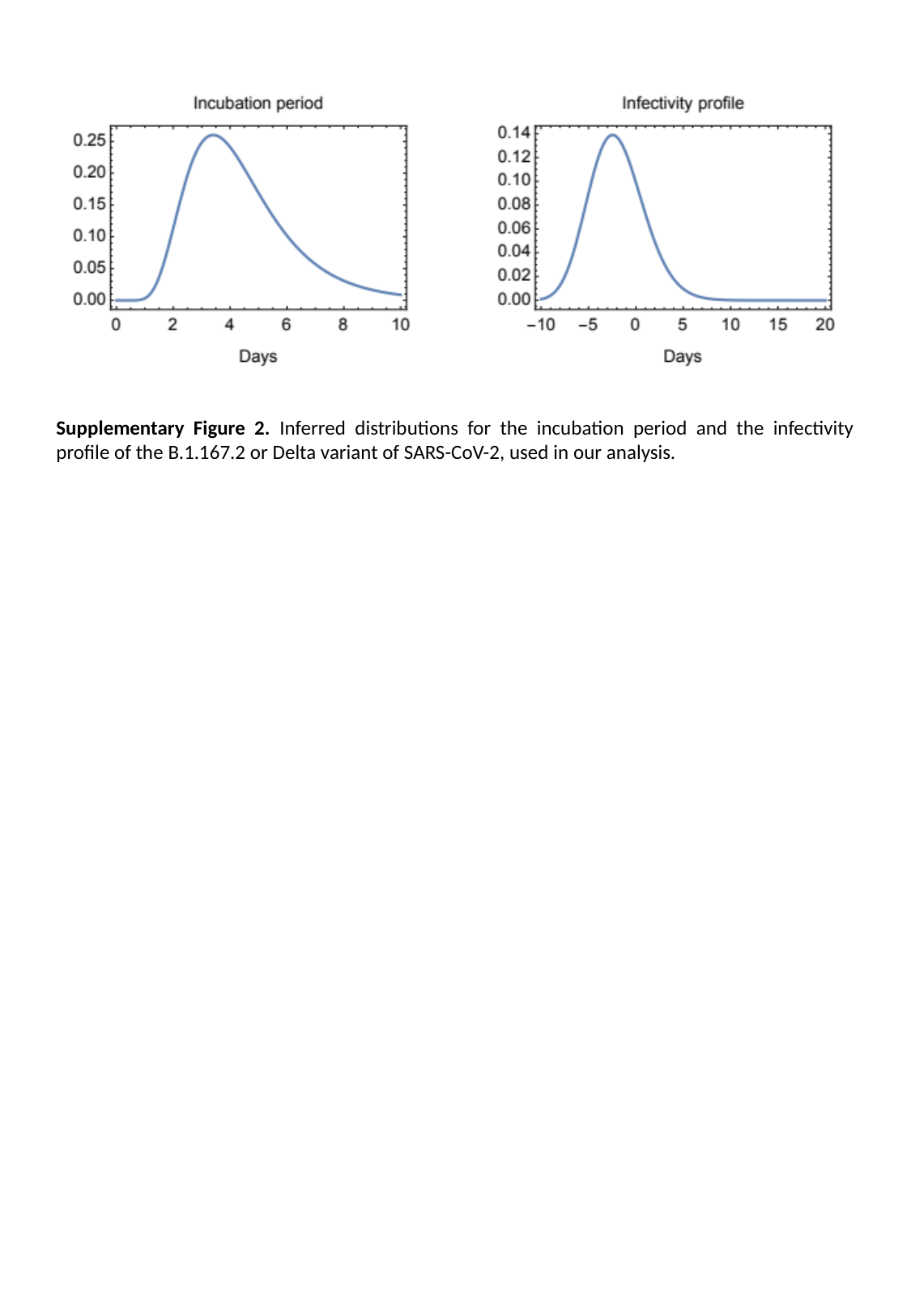

Supplementary Figure 2. Inferred distributions for the incubation period and the infectivity profile of the B.1.167.2 or Delta variant of SARS-CoV-2, used in our analysis.

### Slide 3
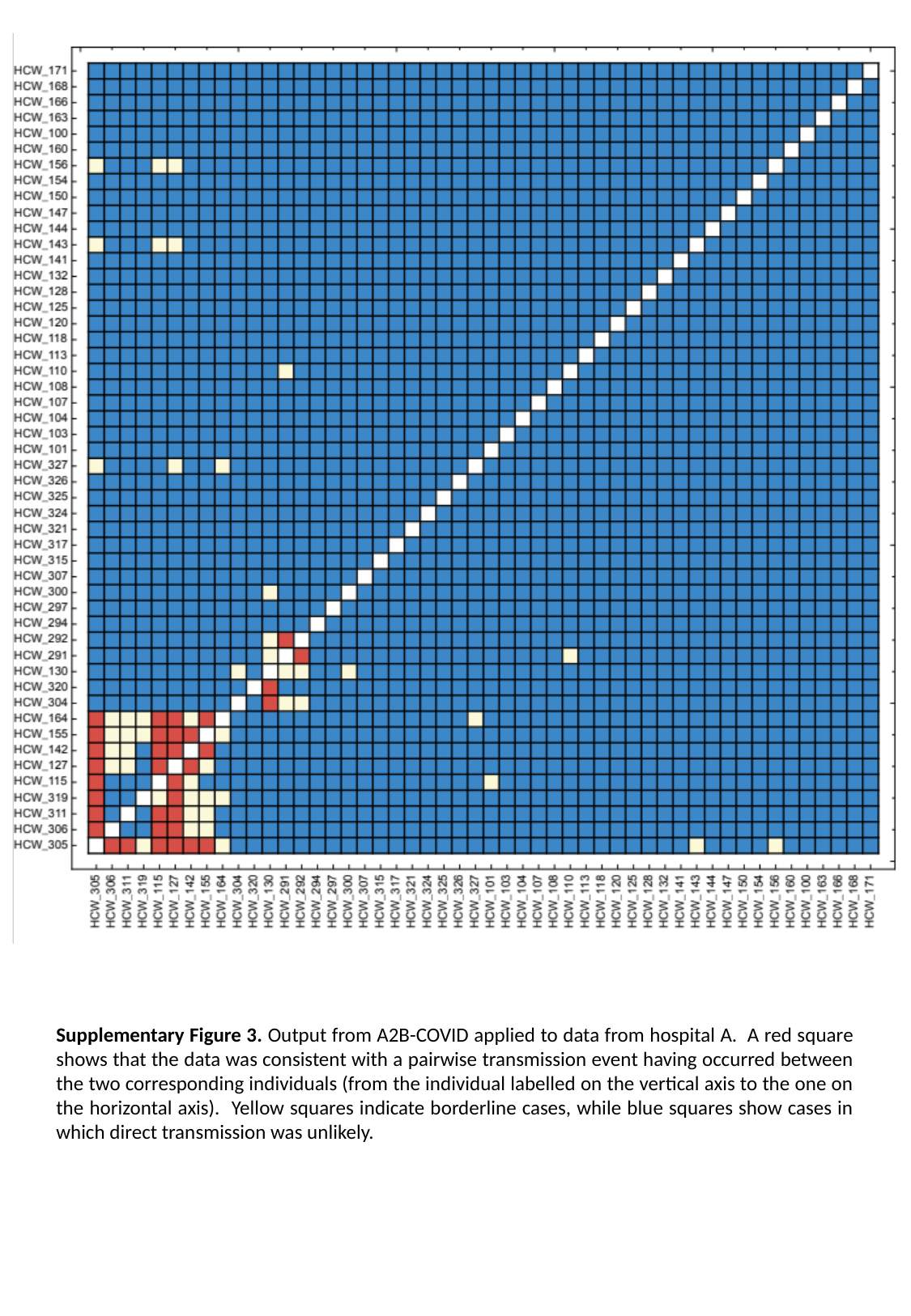

Supplementary Figure 3. Output from A2B-COVID applied to data from hospital A. A red square shows that the data was consistent with a pairwise transmission event having occurred between the two corresponding individuals (from the individual labelled on the vertical axis to the one on the horizontal axis). Yellow squares indicate borderline cases, while blue squares show cases in which direct transmission was unlikely.

### Slide 4
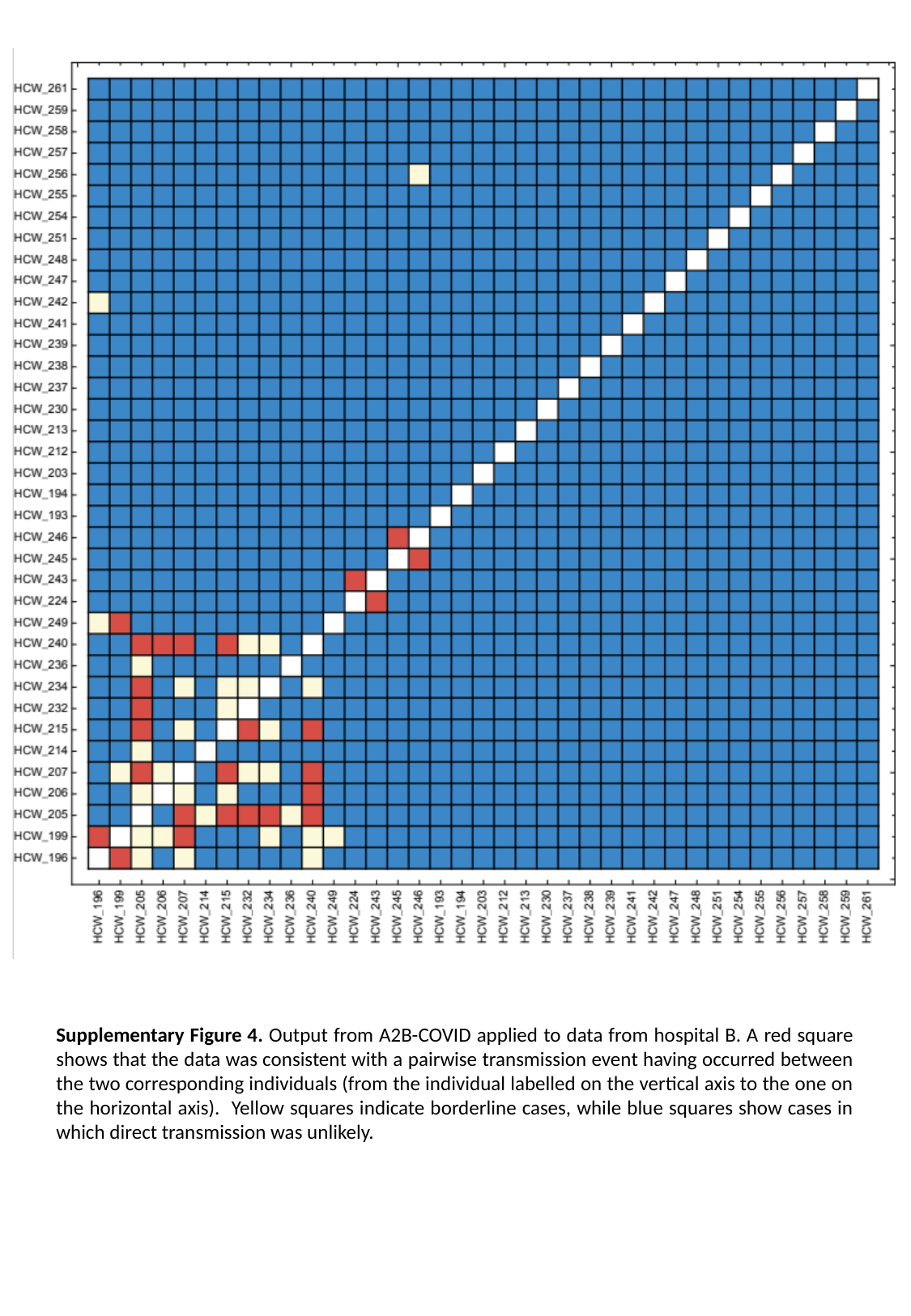

Supplementary Figure 4. Output from A2B-COVID applied to data from hospital B. A red square shows that the data was consistent with a pairwise transmission event having occurred between the two corresponding individuals (from the individual labelled on the vertical axis to the one on the horizontal axis). Yellow squares indicate borderline cases, while blue squares show cases in which direct transmission was unlikely.

### Slide 5
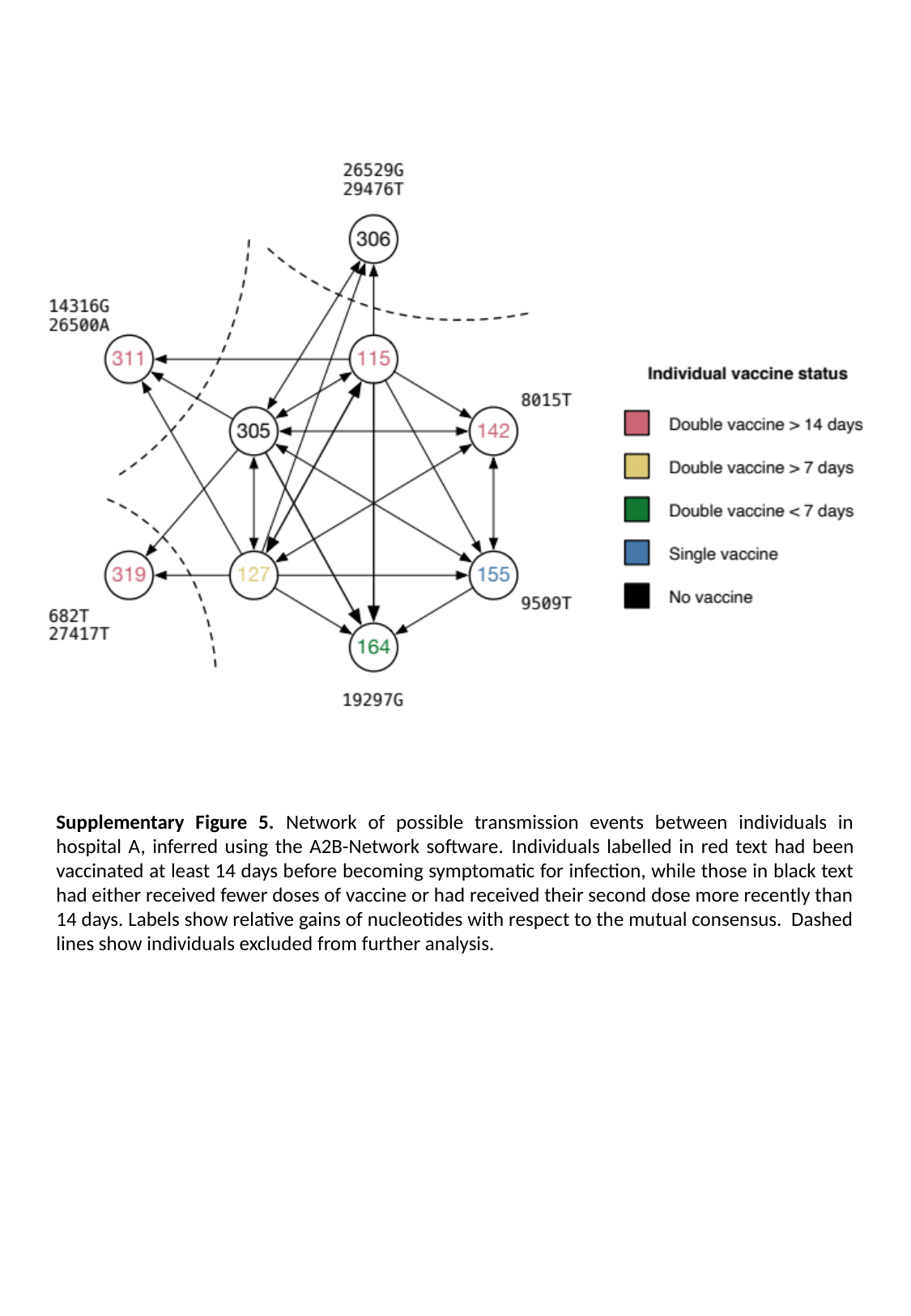

Supplementary Figure 5. Network of possible transmission events between individuals in hospital A, inferred using the A2B-Network software. Individuals labelled in red text had been vaccinated at least 14 days before becoming symptomatic for infection, while those in black text had either received fewer doses of vaccine or had received their second dose more recently than 14 days. Labels show relative gains of nucleotides with respect to the mutual consensus. Dashed lines show individuals excluded from further analysis.

### Slide 6
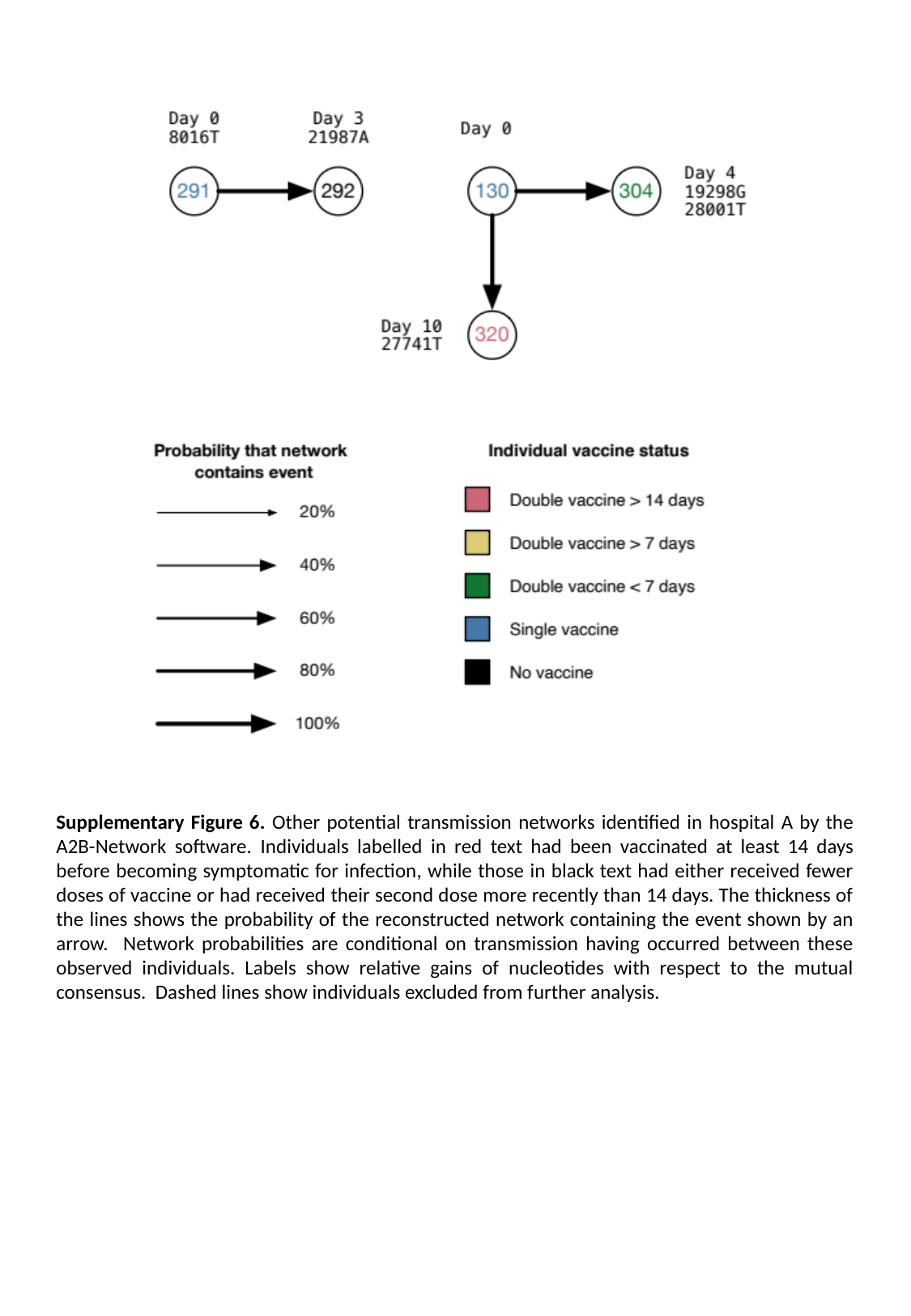

Supplementary Figure 6. Other potential transmission networks identified in hospital A by the A2B-Network software. Individuals labelled in red text had been vaccinated at least 14 days before becoming symptomatic for infection, while those in black text had either received fewer doses of vaccine or had received their second dose more recently than 14 days. The thickness of the lines shows the probability of the reconstructed network containing the event shown by an arrow. Network probabilities are conditional on transmission having occurred between these observed individuals. Labels show relative gains of nucleotides with respect to the mutual consensus. Dashed lines show individuals excluded from further analysis.

### Slide 7
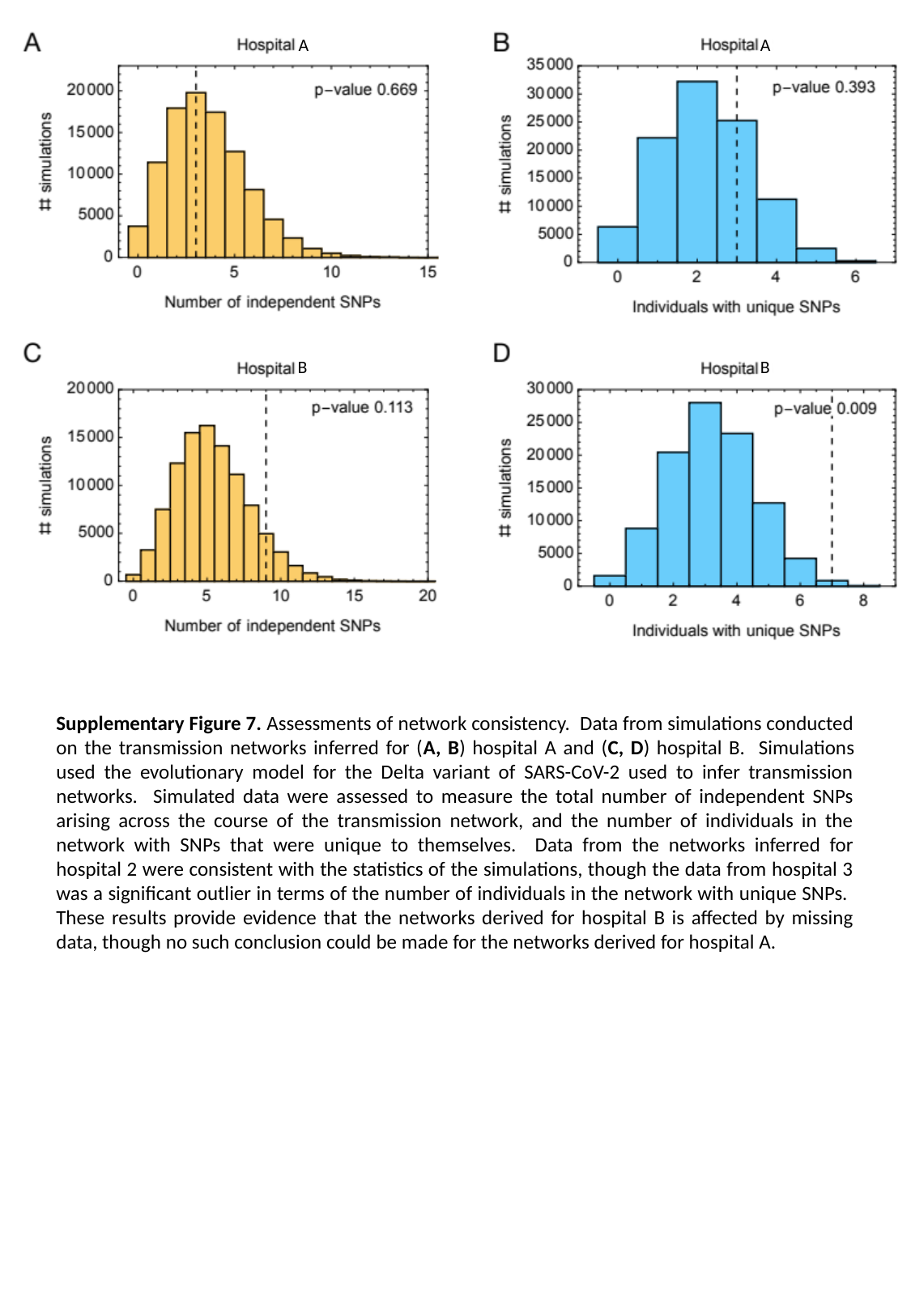

A
A
B
B
Supplementary Figure 7. Assessments of network consistency. Data from simulations conducted on the transmission networks inferred for (A, B) hospital A and (C, D) hospital B. Simulations used the evolutionary model for the Delta variant of SARS-CoV-2 used to infer transmission networks. Simulated data were assessed to measure the total number of independent SNPs arising across the course of the transmission network, and the number of individuals in the network with SNPs that were unique to themselves. Data from the networks inferred for hospital 2 were consistent with the statistics of the simulations, though the data from hospital 3 was a significant outlier in terms of the number of individuals in the network with unique SNPs. These results provide evidence that the networks derived for hospital B is affected by missing data, though no such conclusion could be made for the networks derived for hospital A.

### Slide 8
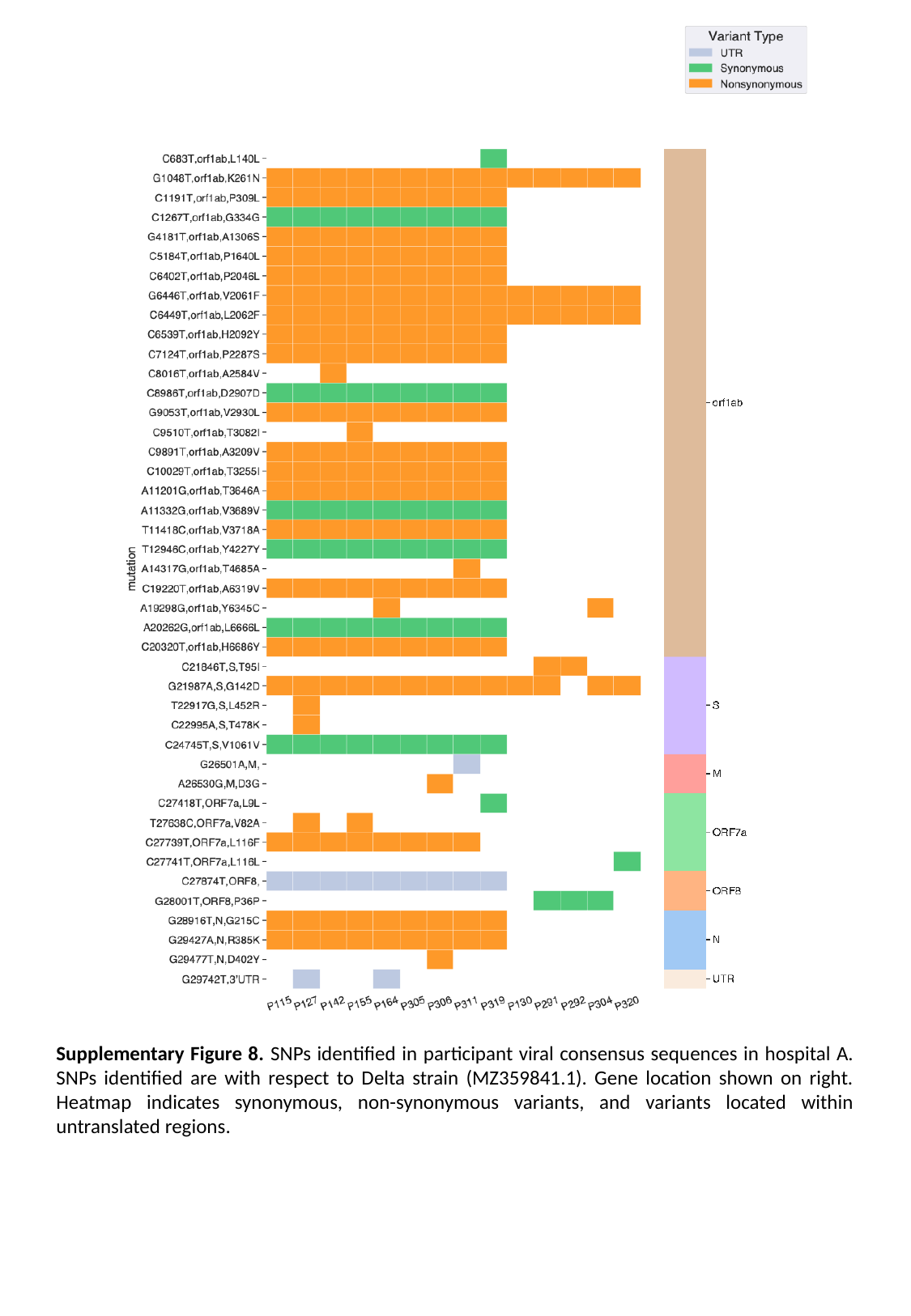

Supplementary Figure 8. SNPs identified in participant viral consensus sequences in hospital A. SNPs identified are with respect to Delta strain (MZ359841.1). Gene location shown on right. Heatmap indicates synonymous, non-synonymous variants, and variants located within untranslated regions.

### Slide 9
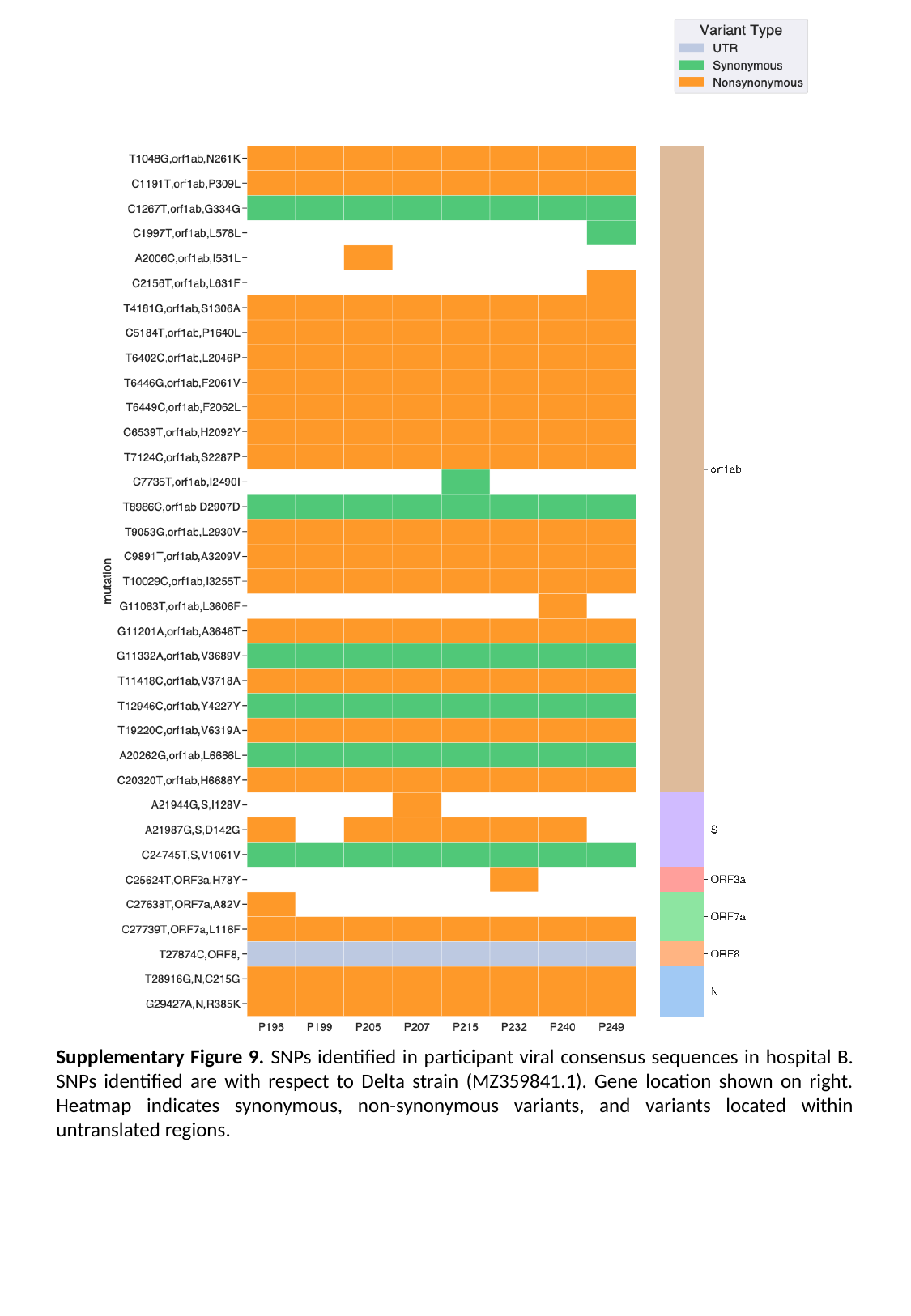

Supplementary Figure 9. SNPs identified in participant viral consensus sequences in hospital B. SNPs identified are with respect to Delta strain (MZ359841.1). Gene location shown on right. Heatmap indicates synonymous, non-synonymous variants, and variants located within untranslated regions.
