## Supplementary material for "Transmission of B.1.617.2 Delta Variant between vaccinated healthcare workers": supp text

**Supporting Information**

**Methods**

*Derivation of statistics for epidemiological modelling*

The A2B-COVID and A2B-Network software packages use two distributions to perform calculations, describing the time from infection to symptom onset, or incubation period of the virus, and the time from symptom onset to infecting another individual, or the infectivity profile of the virus. In the original publication of these packages^1,2^, these parameters were set to model the original strain of the SARS-CoV-2 virus, based upon data and previous calculations described in the literature^3–5^.

In order to evaluate potential transmission events involving the Delta strain of SARS-CoV-2 we used data published on an early study involving this virus^6^ to re-derive parameters for these two distributions.

*Parameters: Incubation period of the virus*

The incubation period of the virus is modelled in A2B-COVID by a lognormal distribution, described as follows:

$$P_{LN}\left( x|\mu,\sigma\right)=\frac{1}{x\sigma\sqrt{2\pi}}exp\left( -\frac{\left( lnx-\mu\right)^{2}}{2\sigma^{2}} \right)$$

Zhang et al report a mean for the incubation period of 4.4 days, with a standard deviation of 1.9 days^6^. We note that the mean and standard deviation of x in the lognormal distribution are given respectively by

$$\bar{x}=exp\left( \mu+\frac{\sigma^{2}}{2} \right)$$

and

$$Std\left( x \right)=\left( exp\left( \sigma^{2} \right)-1 \right)exp\left( 2\mu+\sigma^{2} \right)$$

These equations allow for numerical solution, which was performed using the Mathematica software package, providing the values μ = 1.39599 and σ = 0.41354.

*Parameters: Serial interval*

The serial interval (time between symptom onsets) is modelled by He et al using an offset gamma distribution^3^, with parameters a, b, and o.

$$P_{\gamma}\left( x|a,b,o \right)=\frac{b^{a}\left( x-o \right)^{a-1}exp\left( -b\left( x-o \right) \right)}{\Gamma\left( a \right)}$$

Zhang et al report a mean for this distribution of 2.3 days, with a standard deviation of 3.4 days^6^. We note that the mean and standard deviation of this distribution are given by

$$\bar{x}=ab+o$$

and

$$Std\left( x \right)=a\sqrt{b}$$

These equations allow for straightforward solution. Here the offset is included to account for negative intervals, whereby individual A infects individual B, but B becomes symptomatic before A. Setting the value of the offset o to 20 days (changing this does not greatly affect the shape of the final distribution), the mean and standard deviation are satisfied by the values a = 43.0182 and b = 0.518386.

*Parameters: Infectivity profile*

Following He et al., 2020, we are now in a position to derive an infectivity profile for the Delta strain of SARS-CoV-2, assuming it to follow an offset gamma distribution with parameters α, β, and the offset o=20 days. We used numerical methods to find parameters α and β so as to minimise the distance metric

$$D=\sqrt{\sum_{x=-40}^{30} \left[ P_{\gamma}\left( x|a,b,o \right)-\int_{-\infty}^{\infty} P_{LN}\left( x-y|\mu,\sigma\right)P_{\gamma}\left( y|\alpha,\beta,o \right)dy \right]^{2}}$$

This calculation identifies parameters such that the compound of the time from symptom onset to causing a new infection, and the time from an infection to symptom onset, is as close as possible to the serial interval distribution. From this calculation we obtained the values α = 38.4805 and β = 0.468049.

We note that the results we derive will contain some uncertainty due to the limited data they are based upon, and the precision of the statistics upon which our calculations are based. Our parameters are designed to be broadly representative of SARS-CoV-2 transmission.

**Filtering of data from Hospital 2**

In hospital 2 we restricted our main network calculation to a subset of six cases of HCW infection. A full set of possible transmission events between pairs is shown in Supporting Figure 4, with dashed lines showing the cases removed. We note that the removal of these cases does not affect calculations within the subset of individuals. Specifically, none of the removed cases 306, 311, and 319 can have infected any of the individuals in the subset. Removing 311 and 319 likely reduces the probability that the inferred network involves a case of transmission between HCWs who received their second dose of vaccine 14 days before becoming symptomatic, but none of the removals decrease the value presented in the main text.

**Statistical validation of networks for missing data**

The A2B-Network program performs two distinct calculations on data from a series of infected individuals. Firstly, it carries out a pairwise assessment of potential transmission events, using the approach of A2B-COVID to identify pairs from which the data are consistent with a model of pairwise transmission. We refer to these as plausible transmission events. Secondly, from the set of plausible transmission events, it calculates the likelihoods of potential networks of transmission, linking the infected individuals. Within a network, each individual transmission event must be plausible under the first calculation.

A limitation of the A2B-Network method is that it does not account for missing data. We here used additional statistical testing to evaluate the networks it produced.

Where a network is reconstructed from a dataset in which some samples are missing, the reconstructed transmission tree has the potential to encompass a lesser amount of evolutionary time than the genuine tree. For example, consider an outbreak involving the individuals A, B, C, and D. Suppose that A infects B on day 0, B infects C on day 3, and C infects D on day 6. By day 7, the viruses in the individuals C and D will be separated by eight days of evolution; three days in A, one day in D, and four days in C.

Suppose now that data from the individual B is missing, and a reconstruction is made in which A infects C on day 0, while C infects D on day 6. In this instance, by day 7, the viruses in C and D will be separated by only two days of evolution; one day in each of C and D. While each pairwise transmission event may be plausible, the reconstructed tree will involve less evolutionary time than the genuine tree.

To test whether the transmission networks we inferred, we generated simulations describing virus evolution within these networks, generating statistics to describe the total number of unique mutations observed in the tree, and the number of individuals with mutations that are unique to themselves (i.e. generated in an individual in between the last transmission from that individual to another and the time of sample collection for sequencing).

Given a transmission network, transmission was simulated using the parameters derived for the Delta variant of SARS-CoV-2 above. On day zero, the first infection was initiated, with symptom onset and transmission to other individuals occurring a randomly distributed time after this. The distribution of transmission times was set in a conditional manner such that transmission always occurred after an individual was infected^2^. Genome sequencing was simulated as occurring at the median time after symptom onset of the data from that hospital. A total of 10^5^ simulated networks were generated from each of the statistical ensembles of networks generated by A2B-Network for hospitals 2 and 3.

Network statistics for the data from each hospital are shown in Supplementary Figure 7. While the data from hospital 2 is consistent with the simulated data, the data from hospital 3 is a significant outlier with regards to the number of individuals with unique mutations, providing evidence that the network constructed for hospital 3 is based on incomplete data. While our test does not show that the data from hospital 2 is not incomplete, the inferred network of transmission events is at the least not inconsistent with our underlying evolutionary model.
