## Supplementary material for "Transmission of B.1.617.2 Delta Variant between vaccinated healthcare workers": supp tables

**Supplementary Table 1**: Job roles of vaccinated HCWs in hospitals A and B

| **Job Description** | **Count** |  |
| --- | --- | --- |
| Administrative | 14 | 12.4% |
| Nursing | 36 | 31.9% |
| Medical Physician | 46 | 40.7% |
| Paramedics | 8 | 7.1% |
| Pharmacist | 4 | 3.5% |
| Healthcare assistant | 2 | 1.8% |
| Intern | 2 | 1.8% |
| Laboratory worker | 1 | 0.9% |
| Total | 113 |  |

**Supplementary Table 2:** Lineage assignments of all sequences using the Pangolin tool.

| **Lineage** | **Count** | **Percentage** |
| --- | --- | --- |
| **Alpha (B.1.1.7-like)** | **4** |  |
| B.1.1.7 | 3 | 4.0% |
| Q.1 | 1 | 1.3% |
| **B.1.617.1-like** | **2** |  |
| B.1.617.1 | 2 | 2.7% |
| **Delta (B.1.617.2-like)** | **68** |  |
| B.1.617.2 | 68 | 90.7% |
| **Others** | **1** |  |
| B.1.538 | 1 | 1.3% |
| **Total** | **75** |  |

**Supplementary Table 3:** Symptom prevalence amongst vaccinated HCWs in both hospitals.

| **Symptoms** | Number of cases | % of total |
| --- | --- | --- |
| Asymptomatic | 2 | 1.8% |
| Weakness | 2 | 1.8% |
| Nausea and/or vomiting | 3 | 2.7% |
| Dyspnoea | 3 | 2.7% |
| Congestion | 5 | 4.4% |
| Diarrhoea | 5 | 4.4% |
| Headache | 12 | 10.6% |
| Anosmia and/or ageusia | 16 | 14.2% |
| Sore throat | 32 | 28.3% |
| Myalgia (including backache) | 23 | 20.4% |
| Cough | 49 | 43.4% |
| Fever | 93 | 82.3% |

**Supplementary Table 4:** Details of cases in the inferred transmission networks

| **Individual** | **Job description** | **Symptom onset**  **DD/MM/YY** | **Test date DD/MM/YY** | **CT value** | **Symptoms** |
| --- | --- | --- | --- | --- | --- |
| ***Hospital A*** |  |  |  |  |  |
| P115 | Junior medical staff | 12/04/21 | 13/04/21 | 22.8 | Fever, myalgia, sore throat, abdominal cramps |
| P127 | Nursing student | 14/04/21 | 15/04/21 | 35.2 | Throat irritation |
| P305 | Nursing staff | 17/04/21 | 19/04/21 | 25 | Anosmia,  conjunctivitis, rhinorrhoea |
| P142 | Junior medical staff (ophthalmology) | 18/04/21 | 19/04/21 | 29.6 | Fever, cold cough |
| P155 | Nursing staff | 20/04/21 | 21/04/21 | 29 | Rashes, fever, myalgia, headache |
| P164 | Paramedic | 24/04/21 | 25/04/21 | 21.1 | Fever, cough, sore throat |
| ***Hospital B*** |  |  |  |  |  |
| P232 | Medical officer | 09/04/21 | 12/04/21 | 15.5 | Fever, rhinorrhoea, sore throat |
| P205 | Paediatrician | 12/04/21 | 19/04/21 | 17.1 | Fever, myalgia, anosmia, ageusia |
| P215 | Chief health director / Physician | 13/04/21 | 14/04/21 | 18.5 | Fever |
| P240 | Doctor (pathology labs, COVID wards) | 16/04/21 | 17/04/21 | 12.8 | Fever, cough, myalgia |
| P207 | Physician | 17/04/21 | 19/04/21 | 16.8 | Cough, rhinorrhoea, sore throat |
| P199 | Physician | 22/04/21 | 22/04/21 | 16.3 | Fever, myalgia |
| P196 | OT Assistant | 24/04/21 | 26/04/21 | 14.6 | Fever, sore throat, myalgia |
| P249 | Nursing staff | 28/04/21 | 28/04/21 | 14.6 | Fever, rhinorrhoea |
